# PREVALENCE OF MYOPIA AMONG PUBLIC SCHOOL CHILDREN IN SOUTHERN BRAZIL

**DOI:** 10.1101/2023.05.12.23289894

**Authors:** Patrícia Ioschpe Gus, Raquel Silveira de Maman, Arthur Dementshuk Lengler, Antônia Stumpf Martins, Maria Antônia Torres Arteche, Marina Puerari Pieta, Gabriel Leivas, Rafael Carloto, Diane Marinho, Márcia Beatriz Tartarella, Helena Pakter, Caroline Fabris, Terla Castro, Fernando Kronbauer, Carina Colossi, Monica Jong, Serge Resnikoff

## Abstract

**Purpose:** Myopia has been considered a public health issue by the World Health Organization since 2015. The growing incidence of myopia worldwide, called the myopia epidemic, and its potential blinding complications in adulthood like cataract, glaucoma, retinal detachment and maculopathy, have been extensively published and discussed in peer review papers. Nonetheless, little information about Latin America is available. This study aims to detect the prevalence of myopia in southern Brazil, the biggest country of South America.

**Methods:** A prospective cross sectional study recruited 330 public school children between 2019 and 2021, aged 5 to 20 years old. All children underwent a comprehensive eye examination and detailed lifestyle questionnaire. The Pearson correlation coefficient, Kruskal Wallys and the Chi-Square Test were used to assess simple correlations and associations between myopia and medical conditions, use of medications, ophthalmic history and family history of ocular conditions, besides demographics and lifestyle focused on screen time/day. Associations between the results of the ophthalmologic evaluation and all factors included in the questionnaire were analyzed using the Generalized Estimating Equation model (GEE). The prevalence of hyperopia and astigmatism were also assessed.

**Results:** Total prevalence of myopia was 17.4% (CI 13.8 – 21.7%). Low myopia (-0.50D to -5.75D) comprised 15.2% (CI 11.9 – 19.3%) and high myopia (-6,00D or worse) was 2.1% (CI 1.1 – 4.1%). Relative risk of myopia for females was 1.6 (CI 1.00 – 2.57%) and each additional hour of screen time increased a child’s chance of having myopia by 6.5%. The prevalence of hyperopia was 7,7% (CI 5.4 -10.9%) and of astigmatism, either myopic or hyperopic, was 25.6% (CI 21.4 – 30.2%).

**Conclusions:** Brazil has always been considered a hyperopic country. These are the highest reported prevalences of myopia under cycloplegia and the first paper to present myopia as a more prevalent refractive error than hyperopia among Brazilian school children to date.

## Introduction

Myopia was recognized as a public health issue by the WHO in 2015 and is known to significantly increase the risk of complications and morbidity in the fourth decade of life, such as cataract, glaucoma, retinal detachment and myopic maculopathy [1-4]. East Asia has the highest prevalence of myopia in the world, and, alongside this, a growing prevalence of sight threatening myopia related pathologies [2,4,5]. Myopia is increasing worldwide and population-based eye health data at the country level is needed to develop strategies for public health interventions [3,6,5].

Japan, China, Singapore, South Korea and Taiwan report prevalences of 80% or more young adults with myopia [7,8,9]. Countries of Europe and the United States have myopia rates of 30.6% and 25.4%, respectively, which rises to almost 50% if only the young population is considered (25 -29 years old) [7,10]. Risk factors include high demanding education, many hours of reading and near work daily, little time of activities outdoors, urban areas lifestyle, parental high level education, female sex and ethnicity (Asians are mostly affected) [1-9]. More recently, due to the Covid-19 lockdowns, the incidence of myopia has increased approximately 4% in Chinese children aged 3 to 6 years [10]. The prevalence of myopia is lower in middle and low-income countries are described as ranging from 1.4 to 14.4% in Latin America and 3.4 to 11.4% in Africa, based on ancient statistics [11,12].

There is little information about myopia in Brazil. Local studies have used different methodology, examined different populations and did not all use cycloplegia, which tends to overestimate myopia prevalence [13-17]. Vilar et al. compared two studies conducted in the same ophthalmology hospital in Goiânia (Goiás) at different periods of time [16]. In the evaluation carried out between 1995 and 2000, the prevalence of myopia was 3.6%; in 2014, the study found the prevalence of myopia was 9% [16]. Garcia et al. reported the prevalence of myopia among all refractive errors in Northeastern Brazil in 2001 was 13.3% in randomly selected students [14].

A recent review paper reported that the prevalence of myopia in Brazil was 3.6 to 9.6%, but it was based on publications from several decades ago [12]. Recent papers describe higher prevalences, with rates that varied between 15.2 and 20.4% [13,17]. The first, a retrospective study carried out in São Paulo, evaluated school children under cycloplegia [17]. The second, a prospective cross-sectional study carried out in the equatorial region, evaluated school children under no cycloplegia [13]. The prevalence of myopia in southern Brazil has never been studied. There is a need to better understand the prevalence of myopia in Brazil to develop public health strategies.

## Materials and Methods

This cross-sectional study was conducted in children from public schools in the region of Porto Alegre, Rio Grande do Sul. Hospital de Clínicas de Porto Alegre Ethics Committee approved this study, which adhered to the tenets of the Declaration of Helsinki. Guardians of the children provided written informed consent. All children underwent a comprehensive ophthalmologic examination. The children were not previously screened. A questionnaire on sociodemographic characteristics was completed by participants.

### Sampling

Participants were public school children in the region of Porto Alegre and the study was part of a charitable program that provided spectacles for refractive disorders. Schools were selected by the Public Ministry. Considering the prevalence of myopia in Brazilian school children as 20.4% based on a recent study [13], a sample size of 250 was needed to achieve a 95% confidence level and a confidence interval of 10% [18,19]. The final number of participants determined was 278 to allow for up to 10% non participation [19].

The study began in 2019, was paused due to the novel coronavirus pandemic and resumed in 2021. Children included in the study were aged 5 to 20 years. Exclusion criteria was severe neuro psychomotor developmental disorders who were unable to report visual acuity, subjects with congenital eye disorders (cataract, anophthalmic cavity, strabismus and glaucoma) and contact lens wear on the date of the exam. The examinations were conducted by medical residents and staff using a standardized protocol at 5 different medical centers in Porto Alegre: Hospital de Clínicas de Porto Alegre, Hospital Nossa Senhora da Conceição, Complexo Santa Casa, Centro de Olhos Rio Grande do Sul and Instituto Ivo Corrêa Meyer. A questionnaire on sociodemographic characteristics was completed by participants.

### Defintions

Myopia was defined as a cycloplegic spherical equivalent refraction equal to or worse than -0.50D [20]. The cut off for high myopia was defined as -6,00D or worse [10]. Spherical equivalents greater than or equal to +2.00D were defined as hyperopia [21,22]. Astigmatism was defined as -1.00D (cylindrical diopters) or more [21,23].

### Measurements

All students were at first submitted to a non cycloplegic auto-refraction of both eyes (HRK 7000 Huvitz, South Korea), with at least 3 measurements per eye. Binocular UCVA and BCVA for distance were determined using a standard Snellen chart. School children with UCVA of 0 LogMAR without complaints did not undergo cycloplegia. School children with visual acuity of 0.10 LogMAR or worse, or 0 LogMAR with visual complaints, were instilled with 1% tropicamide (1 drop in each eye, repeated 5 minutes later) and evaluated after 25-30 minutes, with autorefraction (3 measurements per eye), followed by binocular subjective refraction. In hyperopic patients, binocular subjective dynamic refraction was performed prior to pupil dilation with mydriatic eye drops. All participants underwent corneal tomography using Galilei G4 (Ziemer, Germany) for keratometry and corneal thickness measurements. Keratometric keratoconus was defined as maximum keratometry >47.2D [24]. Ocular biometry using ultrasonic AL-100 biometer (Tomey, Japan) was used for axial length measurement, 3 measurements per eye. Subsequent evaluation consisted of retinal mapping and slit lamp biomicroscopy.

A questionnaire about lifestyle, medical and family history for ocular and systemic diseases, itchy eyes, history of asthma, bronchitis or rhinitis, hours of daily screen time (including cell phone, tablet and computer) and medications in use was completed by the participants and their guardians.

### Statistics

All statistical analysis was performed using SPSS version 18.0 software (SPSS, Chicago). The Pearson Correlation Coefficient, Kruskal Wallys and the Chi-Square Test were used to assess simple correlations and associations between variables. Associations between the results of the ophthalmologic evaluation and all factors included in the questionnaire were analyzed using the Generalized Estimating Equation Model (GEE). Confidence interval of 95% and p ≤ 0.05 were chosen for statistical significance. Association between demographics and lifestyle with low and high myopia were assessed too.

## Results

### Participants

A total of 333 school children were evaluated in this study and 52% of those were males. Ethnicity based on self classification was 37.5% white, 39% afro-descendants and 0.4% other; 23.1% did not classify themselves. Mean age was 12.74 years (95% CI 12.38-13.10). More than half were aged 10-15 years (53.4%); 20.4% were younger than 9 years and 26.1% were older than or equal to 16 years. Most participants were from the Hospital de Clínicas de Porto Alegre (52%). Three children were excluded from the analysis due to technical difficulties during the measurements (Table 1).

**Table 1.** Demographics of the 333 study participants

|  | N° | Rate (%) |
| --- | --- | --- |
| **Sex** |  |  |
| Male | 173 | 52 |
| Female | 160 | 48 |
| **Skin Color** |  |  |
| White | 125 | 37.5 |
| Afro-descendant | 131 | 39 |
| Not Referred | 77 | 23.1 |
| **Age** |  |  |
| <= 9 | 68 | 20.4 |
| 10 - 15 | 178 | 53.4 |
| >=16 | 87 | 26.1 |
| Medical Center |  |  |
| HCPA | 174 | 52 |
| HNSC | 40 | 12 |
| CSC | 39 | 12 |
| CORS | 43 | 13 |
| IICM | 37 | 11 |

Note: HCPA = Hospital de Clínicas de Porto Alegre; HNSC = Hospital Nossa Senhora da Conceição; CSC = Complexo Santa Casa; CORS = Centro de Olhos Rio Grande do Sul; IICM = Instituto Ivo Corrêa Meyer

Approximately 51% of children achieved UCVA 0 LogMar bilaterally and 34.5% wore spectacles. Average screen time use was 4.92 hours (95% CI 4.48-5.35). 63.4% of children rubbed their eyes; 28% used chronic medications for non-ocular conditions; 26% had some type of allergy or atopy (allergic conjunctivitis; rhinitis or asthma), but only 4% used eye drops (antihistamine and/ or lubricants) regularly.

### Prevalence of refractive errors

Prevalence of myopia was 17.4% (CI 13.8 – 21.7%). Low myopia corresponded to 15.2% (CI 11.9 – 19.3%), while high myopia was present in 2.1% (CI 1.1 – 4.1%). Mean myopia was -2.73 (SD 2.69). Hyperopia prevalence was 7.7% (CI 5.4 -10.9%) and astigmatism prevalence was 25.6% (CI 21.4 – 30.2%)(Table 2).

**Table 2.** Refractive errors prevalence

|  | N | Rate (%) | 95% CI |
| --- | --- | --- | --- |
| Myopia ($\geq -0.50D$) | 58 | 17.4 | 13.8 - 21.7 |
| High Myopia ($\geq -6.00D$) | 7 | 2.1 | 1.1-4.1 |
| Low Myopia (-0.50 to -5.75D) | 51 | 15.2 | 11.9-19.3 |
| Hyperopia ($\geq +2.00D$) | 26 | 7.7% | 5.4-10.9 |
| Astigmatism ($\leq 1.00D$) | 1.06 | 25.6% | 21.4-30.2 |

Note: CI = Confidence Interval

### Risk factors associated with myopia

Sex disclosed a significant difference in myopia prevalence: 21.3% for females and 13.2% for males (p<0.01). Females presented a relative risk of 1.6 (CI 1.00 – 2.57%) (p=0.047). Each additional hour of screen time increased a child’s chance of having myopia by 6.5% (CI 1.01-1.12%) (*p*=0.01). No association was found between ethnicity, age or rubbing the eyes with low or high myopia. Table 3 shows the association between risk factors and myopia.

**Table 3.** Association between risk factors and myopia

|  | RR | 95% CI | SD | *p* |
| --- | --- | --- | --- | --- |
| Sex | 1.6 | [1.00-2.57] | - | 0.04 |
| Age Groups | - | - | 0,380 | 0.18 |
| Ethnicity | - | - | 0,385 | 0.367 |
| Rubbing the eyes | 0.63 | [0.40-0.98] | - | 0.043 |
| Screen Time Use | 1.06 | [1.02-1.12] | - | 0.01 |

*Note.* RR = Relative Risk; CI = Confidence Interval; SD = Standard Deviation

Considering the cut-off of -6.00D for high myopia, maximum corneal power differed between low, with 45.1D (SD 2.00), and high myopia, with 45.7D (SD 3.00)(*p*<0.01). Axial length also showed statistically significant differences between low and high myopia: 23.58mm (SD 1.03) and 26.62mm (SD 1.01), respectively, with p<0.01. However, we found no statistically significant difference in keratometric keratoconus between groups (p=0.56). Central corneal thickness was not significantly different between low and high myopia: 532 microns (SD 33) for low and 538 microns (SD 56) for high myopia (p=0.84).

## Discussion

There is limited information regarding the prevalence of myopia in Brazil and this is the first study on myopia prevalence in southern Brazil [13,17]. Papers published previously in other regions used different methodology (no cycloplegia; retrospective analysis; adults included; patients from ophthalmology centers) [11,15,16]. The data collected in Porto Alegre is representative of public school children from the entire country in terms of the ethnic mix, since the proportion of Caucasians and Afro-descendants is similar to the national distribution[25]. Prevalence of myopia between Caucasians and Afro-Descendants were not significantly different [9,26-28].

Environmental risk factors are known to be important in myopia development [29]. An important difference between the south and the equatorial regions of Brazil is the weather. It is hot and sunny in the north and northeast for the entire year, while the south is subtropical and has four seasons. We found mean myopia was higher in the south compared to the equatorial Brazil (-2.73D in the south and -0,50D in the northeast), although both are still low [13]. Nevertheless, the difference in temperature and consequently different time of exposure to sunlight per week might influence further development of myopia in the different regions [5,13,28,30-33].

High levels of education are known to strongly relate to myopia [29]. Public schools in Brazil generally have unfortunately weak academic outcomes and are not demanding [35-36]. Public schools tend to have one four hour shift of classes per day with no demanding homeworks, and had no activities during the Novel Coronavirus Pandemic [38]. Nevertheless, all social classes have access to screens and the children spend most of the time indoors, using near work electronics during their spare time [39]. This might explain the apparent growing prevalence of myopia over the last decade among Brazilian school children and the higher risk of 6.5% for additional hours of near work activities reported by this study [40,41].

No private school children were included to avoid a possible bias, since they had access to education online during the pandemic and tend to have better quality of education overall, with more hours of schooling per day and more demanding homeworks [42]. Private school children represent a small proportion of students in Brazil: 17% [36,43]. The authors plan to perform another study including private schools.

Exclusion of keratoconus based on careful evaluation of the corneas was performed. Keratoconus is a relatively frequent progressive bilateral ectasia that might cause blindness in adolescents and young adults, which also presents with myopia and astigmatism and might act as a confounding factor. Although keratometric keratoconus and central corneal thickness did not differ between myopia and non myopia groups in this study, rubbing the eyes increases the risk for keratoconus development [47-51] and many school children complained of itchy eyes. They were treated for different allergies (rhinitis and asthma) with oral drugs, but not with adequate eye drops.

Itchy eyes may also be caused by chronic dry eyes associated with excessive near work activities [40,52-56]. Proactively asking about rubbing the eyes during an ophthalmologic evaluation and informing parents and children about the risks and causes of eye rubbing should be considered as part of the eye examination routine. Besides, prescribing lubricants and antihistamine eye drops in cases of dry eyes and mild allergic conjunctivitis in children is advisable.

Most school children had excellent uncorrected visual acuity and the prevalence of myopia is still low in comparison to other continents, but it seems to be consistently increasing overtime [13-17]. There was an increased risk of myopia in females, previously also described in the literature [45,46]. The main possible bias of this study is the non-randomly selected sample. However, no children had previously had a vision screening and the eye exam was comprehensive with objective and subjective refraction done under cycloplegia, which performs a complete and precise evaluation.

Further studies understanding the prevalence of myopia in other parts of Brazil and other Latin American countries, and the factors associated with it are needed. Public health initiatives that aim to prevent myopia from increasing further and protect our future adult generations from vision permanent impairment must be taken into consideration.

## Data Availability

All relevant data are within the manuscript and its Supporting Information files.

## Acknowledgements

The authors acknowledge Hospital de Clínicas Central Administration and Outpatient Coordination, the employees and staff members of Ophthalmology of Hospital de Clínicas de Porto Alegre, the financial support of FIPE (Fundo de Incentivo à Pesquisa do Hospital de Clínicas de Porto Alegre), the Biostatistics of GPPG (Grupo de Pesquisa e Pós-Graduação do Hospital de Clínicas de Porto Alegre), the medical students, residents and fellows of Ophthalmology from all facilities, the Public Ministry of Porto Alegre and Viamão, Sorigs (Sociedade de Oftalmologia do Rio Grande do Sul) and the International Myopia Institute. Their support was essential for collecting data during the Coronavirus Pandemic and for helping to disseminate Brazilian myopia prevalence to the world in the International Myopia Conference 2022.

## Authors Contributions

**Patricia Ioschpe:** Conceptualization (lead); data curation (lead); formal analysis (lead); investigation (lead); methodology (lead); project administration (lead); supervision (lead); writing – original draft (lead); writing – review and editing (lead). **Raquel de Maman**: data curation (equal); formal analysis (equal); writing – original draft (equal). **Arthur Lengler:** data curation (equal); formal analysis (equal); writing – original draft (equal). **Maria Antônia Arteche**: data curation (equal), **Marina Pieta**: data curation (equal). **Gabriel Leivas**: data curation (equal); formal analysis (equal).

**Rafael Carloto**: data curation (equal); formal analysis (equal). **Diane Marinho**: Conceptualization (supporting); data curation (equal). **Márcia Tartarella**: Conceptualization (supporting). **Helena Pakter:** data curation (supporting), **Caroline Fabris**: data curation (supporting). **Terla Castro**: data curation (supporting). **Fernando Kronbauer:** data curation (supporting). **Carina Colossi:** data curation (supporting). **Monica Jong**: writing – review and editing (supporting). **Serge Resnikoff** writing – review and editing (supporting).

## Bibliography

1. The impact of myopia and high myopia: report of the Joint World Health Organization – Brien Holden Vision Institute [Internet]. Global Scientific Meeting on Myopia, University of New South Wales, Sydney, Australia, 16–18 March 2015. 2 [cited 2022 May 21]. Available from: https://myopiainstitute.org/wp-content/uploads/2020/10/Myopia_report_020517.pdf.

2. Németh J, Tapasztó B, Aclimandos WA, et al. Update and guidance on management of myopia. European Society of Ophthalmology in cooperation with International Myopia Institute. Eur J Ophthalmol. 2021 May;31(3):853–883.

3. Holden BA, Fricke TR, Wilson DA, et al. Global Prevalence of Myopia and High Myopia and Temporal Trends from 2000 through 2050. Ophthalmology. 2016 May;123(5):1036–42.

4. Ohno-Matsui K, Lai TY, Lai CC, Cheung CM. Updates of pathologic myopia. Prog Retin Eye Res. 2016 May;52:156–87.

5. Sankaridurg P, Tahhan N, Kandel H, et al. Impact of Myopia. Invest Ophthalmol Vis Sci. 2021 Apr 28;62(5):2

6. Dolgin E. The myopia boom. Nature. 2015 Mar 19;519(7543):276–8.

7. Recko M, Stahl ED. Childhood myopia: epidemiology, risk factors, and prevention. Mo Med. 2015 Mar-Apr;112(2):116–21.

8. Morgan IG, French AN, Ashby RS, Guo X, Ding X, He M, Rose KA. The epidemics of myopia: Aetiology and prevention. Prog Retin Eye Res. 2018 Jan;62:134–149.

9. Wu PC, Huang HM, Yu HJ, Fang PC, Chen CT. Epidemiology of Myopia. Asia Pac J Ophthalmol (Phila). 2016 Nov/Dec;5(6):386–393.

10. Wolffsohn JS, Flitcroft DI, Gifford KL, et al. IMI - Myopia Control Reports Overview and Introduction. Invest Ophthalmol Vis Sci. 2019 Feb 28;60(3):M1–M19.

11. Grzybowski A, Kanclerz P, Tsubota K, Lanca C, Saw SM. A review on the epidemiology of myopia in school children worldwide. BMC Ophthalmol. 2020 Jan 14;20(1):27.

12. Ang, Saini. Updates on Myopia. Springer, Singapore; 2020. p 27–48.

13. Yotsukura E, Torii H, Ozawa H, et al. Axial Length and Prevalence of Myopia among Schoolchildren in the Equatorial Region of Brazil. J Clin Med. 2020 Dec 31;10(1):115.

14. Lira RP, Arieta CE, Passos TH, el al.Distribution of Ocular Component Measures and Refraction in Brazilian School Children. Ophthalmic Epidemiol. 2017 Feb;24(1):29–35.

15. Garcia CA, Oréfice F, Nobre GF, Souza Dde B, Rocha ML, Vianna RN. Prevalence of refractive errors in students in Northeastern Brazil. Arq Bras Oftalmol. 2005 May-Jun;68(3):321–5.

16. Vilar MC, Abrahão MM, Mendanha Dde A, et al. Increased prevalence of myopia in an ophthalmologic hospital in Goiânia – Goiás. Rev Bras Oftalmol. 2016 sep-oct; 75(5): 356–9. https://doi.org/10.5935/0034-7280.20160071

17. Costa DR, Debert I, Sussana FN, Falabreti JG, Polati M, Júnior RS. Vision for the Future Project: Screening impact on the prevention and treatment of visual impairments in public school children in São Paulo City, Brazil. Clinics. 2021;76: e3062 https://doi.org/10.6061/clinics/2021/e3062

18. Yang M, Luensmann D, Fonn D, et al. Myopia prevalence in Canadian school children: a pilot study. Eye (Lond). 2018 Jun;32(6):1042–1047.

19. Borges R, Mancuso A, Camey S, et al. Power and Sample Size for Health Researchers: a tool for calculating sample size and statistical power designed for health researchers. Clin. biomed. res. 2021; 40(4): 247–253, 2020. ilus.

20. Flitcroft DI, He M, Jonas JB, et al. IMI - Defining and Classifying Myopia: A Proposed Set of Standards for Clinical and Epidemiologic Studies. Invest Ophthalmol Vis Sci. 2019 Feb 28;60(3):M20–M30.

21. Harrington SC, Stack J, Saunders K, O’Dwyer V. Refractive error and visual impairment in Ireland schoolchildren. Br J Ophthalmol. 2019 Aug;103(8):1112–1118.

22. Castagno VD, Fassa AG, Carret ML, Vilela MA, Meucci RD. Hyperopia: a meta-analysis of prevalence and a review of associated factors among schoolaged children. BMC Ophthalmol. 2014 Dec 23;14:163.

23. O’Donoghue L, Breslin KM, Saunders KJ. The Changing Profile of Astigmatism in Childhood: The NICER Study. Invest Ophthalmol Vis Sci. 2015 May;56(5):2917–25.

24. Belin MW, Jang HS, Borgstrom M. Keratoconus: Diagnosis and Staging. Cornea. 2022 Jan 1;41(1):1–11.

25. Instituto Brasileiro de Geografia e Estatística (IBGE) [Internet] 2012-2019. [updated 2022; cited 2022 Jan 24]. Available from: https://educa.ibge.gov.br/jovens/conheca-o-brasil/populacao/18319-cor-ou-raca.html

26. Pan CW, Ramamurthy D, Saw SM. Worldwide prevalence and risk factors for myopia. Ophthalmic Physiol Opt. 2012 Jan;32(1):3–16.

27. Morgan IG, French AN, Ashby RS, et al. The epidemics of myopia: Aetiology and prevention. Prog Retin Eye Res. 2018 Jan;62:134–149.

28. He M, Xiang F, Zeng Y, et al. Effect of Time Spent Outdoors at School on the Development of Myopia Among Children in China: A Randomized Clinical Trial. JAMA. 2015 Sep 15;314(11):1142–8.

29. Wolffsohn JS, Jong M, Smith EL 3r, et al. IMI 2021 Reports and Digest - Reflections on the Implications for Clinical Practice. Invest Ophthalmol Vis Sci. 2021 Apr 28;62(5):1. doi: 10.1167/iovs.62.5.1. PMID: 33909037; PMCID: PMC8083124.

30. Ramamurthy D, Lin Chua SY, Saw SM. A review of environmental risk factors for myopia during early life, childhood and adolescence. Clin Exp Optom. 2015 Nov;98(6):497–506.

31. Bremond-Gignac D. Myopie de l’enfant [Myopia in children]. Med Sci (Paris). 2020 Aug-Sep;36(8-9):763–768.

32. Cooper J, Tkatchenko AV. A Review of Current Concepts of the Etiology and Treatment of Myopia. Eye Contact Lens. 2018 Jul;44(4):231–247.

33. Jones-Jordan LA, Sinnott LT, Chu RH, et al. Myopia Progression as a Function of Sex, Age, and Ethnicity. Invest Ophthalmol Vis Sci. 2021 Aug 2;62(10):36.

34. Mountjoy E, Davies NM, Plotnikov D,et al. Education and myopia: assessing the direction of causality by mendelian randomisation. BMJ. 2018 Jun 6;361:k2022. doi: 10.1136/bmj.k2022. Erratum in: BMJ. 2018 Jul 4;362:k2932.

35. Education Policy Outlook Brasil [Internet] 2022. [updated 2022, cited 2022 jun 22]. Available from:https://www.oecd.org/education/policy-outlook/country-profile-Brazil-2021-INT-PT.pdf (pg 35–40)

36. Portal da Transparência Brasil [Internet] 2022. [updated 2022; cited 2022 jun 22]. Available from: https://www.portaltransparencia.gov.br/funcoes/12-educacao?ano=2022

37. Associação Brasileira de Recursos Humanos (ABRH) 2017 [Internet]: [cited 2022 jun 22]. Available from: http://www.abrhce.com.br/brasil-ocupa-60a-posicao-em-ranking-de-educacao-em-lista-com-76-paises.

38. Ministério da Educação (MEC) [Internet] [updated 2022, cited 2022 jun 22]. Available from: http://portal.mec.gov.br/component/tags/tag/jornada-escolar

39. Instituto Brasileiro de Geografia e Estatística (IBGE) [Internet] 2018-2019. [updated 2020; cited 2022 jun 22]. Available from: https://educa.ibge.gov.br/jovens/materias-especiais/20787-uso-de-internet-televisao-e-celular-no-brasil.html

40. Huang HM, Chang DS, Wu PC. The Association between Near Work Activities and Myopia in Children-A Systematic Review and Meta-Analysis. PLoS One. 2015 Oct 20;10(10):e0140419.

41. Agência Brasil [Internet] 2018-2019. [updated 2020; cited 2022 jun 22]. Available from:https://agenciabrasil.ebc.com.br/educacao/noticia/2021-04/mais-de-5-milhoes-de-criancas-e-adolescentes-ficaram-sem-aulas-em-2020

42. Sampaio B, Guimarães J. Diferenças entre ensino público e privado no Brasil. Econ. Apl. 2009 May; 13 (1).

43. Ministério da Educação (MEC) Censo Escolar 2021 [Internet]. [updated 2022; cited 2022 jun 22]. Available from: https://download.inep.gov.br/censo_escolar/resultados/2021/apresentacao_coletiva.pdf.

44. Yotsukura E, Torii H, Inokuchi M, et al. Prevalence of Myopia and Association of Myopia With Environmental Factors Among Schoolchildren in Japan. JAMA Ophthalmol. 2019 Nov 1;137(11):1233–1239.

45. Rudnicka AR, Kapetanakis VV, Wathern AK, et al. Global variations and time trends in the prevalence of childhood myopia, a systematic review and quantitative meta-analysis: implications for aetiology and early prevention. Br J Ophthalmol. 2016 Jul;100(7):882–890.

46. Ludwig CA, Boucher N, Saroj N, Moshfeghi DM. Differences in anterior peripheral pathologic myopia and macular pathologic myopia by age and gender. Graefes Arch Clin Exp Ophthalmol. 2021 Nov;259(11):3511–3513.

47. Sahebjada S, Al-Mahrouqi HH, Moshegov S, et al. Eye rubbing in the aetiology of keratoconus: a systematic review and meta-analysis. Graefes Arch Clin Exp Ophthalmol. 2021 Aug;259(8):2057–2067.

48. Najmi H, Mobarki Y, Mania K, et al. The correlation between keratoconus and eye rubbing: a review. Int J Ophthalmol. 2019 Nov 18;12(11):1775–1781.

49. Ahuja P, Dadachanji Z, Shetty R, et al. Relevance of IgE, allergy and eye rubbing in the pathogenesis and management of Keratoconus. Indian J Ophthalmol. 2020 Oct;68(10):2067–2074.

50. Chervenkoff JV, Hawkes E, Ortiz G, Horney D, Nanavaty MA. A randomized, fellow eye, comparison of keratometry, aberrometry, tear film, axial length and the anterior chamber depth after eye rubbing in non-keratoconic eyes. Eye Vis (Lond). 2017 Aug 14;4:19.

51. Hashemi H, Heydarian S, Hooshmand E. The Prevalence and Risk Factors for Keratoconus: A Systematic Review and Meta-Analysis. Cornea. 2020 Feb;39(2):263–270.

52. Yotsukura E, Torii H, Inokuchi M, et al. Current Prevalence of Myopia and Association of Myopia With Environmental Factors Among Schoolchildren in Japan. JAMA Ophthalmol. 2019 Nov 1;137(11):1233–1239.

53. Karakus S, Mathews PM, Agrawal D, Henrich C, Ramulu PY, Akpek EK. Impact of Dry Eye on Prolonged Reading. Optom Vis Sci. 2018 Dec;95(12):1105–1113.

54. Karakus S, Agrawal D, Hindman HB, Henrich C, Ramulu PY, Akpek EK. Effects of Prolonged Reading on Dry Eye. Ophthalmology. 2018 Oct;125(10):1500–1505.

55. Mathews PM, Ramulu PY, Swenor BS, Utine CA, Rubin GS, Akpek EK. Functional impairment of reading in patients with dry eye. Br J Ophthalmol. 2017 Apr;101(4):481–486.

56. Hom MM, Nguyen AL, Bielory L. Allergic conjunctivitis and dry eye syndrome. Ann Allergy Asthma Immunol. 2012 Mar;108(3):163–6.

